# The effect of natural terrestrial gamma radiation emissions and environmental radon levels on the incidence of cutaneous squamous cell carcinoma of the head and neck

**DOI:** 10.1101/2023.06.27.23291952

**Authors:** Ivan Couto-González, Jorge Arenaz-Búa, Antonio Taboda-Suárez, Abel García-García

**Affiliations:** Plastic and Reconstructive Surgery Department. University Hospital of Santiago de Compostela, Santiago de Compostela, Spain; Oral and Maxillofacial Surgery Department. University Hospital of Santiago de Compostela, Santiago de Compostela, Spain

## Abstract

The effects of ionising radiation on the development of cutaneous squamous cell carcinoma have been previously studied in environments in which the levels of ionising radiation have been increased by artificial sources. The purpose of this study is to determine the role that environmental radon concentration and natural gamma radiation emissions may play in the development of head and neck cutaneous squamous cell carcinoma in a geographical area which is known to have high levels of radon and natural terrestrial gamma radiation emissions. A total of 284 patients diagnosed with cutaneous squamous cell carcinoma during the 26-month observation period were included in the study. The overall incidence was 37.33 cases/100,000 people-year. The mean of environmental radon concentration according to their council of residence was 116.69 Bq/m^3^ (40.05) and the mean of natural terrestrial gamma radiation emitted according to their council of residence was 14.25 μRad/hour (3.86). The multiple linear regression analysis revealed that only mean natural terrestrial gamma radiation emissions in the council of residence (P < 0.05), carrying out an outdoor profession (P < 0.05) and the mean number of hours of sunlight per year in the council of residence (P = 0.03) were found to have statistical significance on the incidence of head and neck cutaneous squamous cell carcinoma. Emissions of natural terrestrial gamma radiation have never been proposed as a factor having an influence on the development of cutaneous neoplasms. The implications that this could have in areas with naturally high terrestrial gamma radiation emissions should be more exhaustively studied to assess the true weight of this factor.

## Introduction

Cutaneous squamous cell carcinoma (cSCC) is the second most frequent non-melanoma skin neoplasm diagnosed worldwide after basal cell carcinoma (BCC). In recent decades, increasing incidence rates of BCC and cSCC have been reported^1^. The effects of ionising radiation on the development of cSCC have been studied in different series. Unlike the general increase in incidence that occurs in BCC, the results in the case of cSCC are more variable^2–4^. These series are usually situated in environments in which the levels of ionising radiation have been increased by artificial sources, usually in workplaces or in the study of the effects caused by the atomic bombs dropped on Japan in the last days of World War II.

It has been demonstrated that natural terrestrial radiation can have an influence on the development of certain neoplasms, such as lung cancer, in environments with high levels of the natural radioisotope ^222^Rn ^5^. Some studies have sought to find a link between environmental exposure to radon and an increase in the incidence of different skin neoplasms with varying results^6–9^. Through the release of alpha particles, radon is responsible for 55% of the ionising radiation naturally received by humans^10^. ^222^Rn particles concentrate a large amount of energy, which, after inhalation of ^222^Rn, could explain the pathogenesis of lung cancer. However, alpha particles have an extremely low capacity of tissue penetration, leading to serious doubts about whether they could have any mutagenic effect on cells in the basal layer of the epidermis, with the exception of areas with thin skin not usually protected by clothing, such as the facial region^11^.

Gamma particles are a type of massless electromagnetic radiation consisting of photons. Their high, massless energy allows them to penetrate matter more deeply than alpha and beta radiation, causing damage to cellular genetic material^12^. The role that natural emissions of gamma radiation from terrestrial emission may play in the development of skin neoplasms has not been studied to date.

The aim of this study is to determine the role that environmental radon concentration and natural gamma radiation emissions may play in the development of head and neck cSCC in a geographical area which is known to have high levels of radon and natural terrestrial gamma radiation emissions. In addition, other factors classically related to the development of cSCC will be analysed.

## Material and Methods

### Patients

All patients diagnosed with head and neck cSCC over a 26-month period ranging from November 2018 to December 2020 in our healthcare area were located through the database of the Anatomical Pathology department. This is the only department capable of carrying out histopathological analyses within the study area. Therefore, the cases processed correspond to the total cases diagnosed. The inclusion criterion was the diagnosis of head and neck cSCC during the period of study. Patients already included in the database who presented with a second head and neck cSCC during the study period were excluded. In the case of synchronous cSCC, the most severe tumour was always that included in the study according to the criteria of the presence of regional lymph node or distant metastasis, infiltration of local structures, tumour differentiation, tumour diameter and infiltration depth. Patients with partial follow-up in other centres for whom it was not possible to gain complete access to their clinical history were also excluded from the study.

### Study protocol

A study protocol was developed taking into account demographic, occupational and personal history factors of the patients which could have a known influence on the development of cSCC. Clinical and histopathological data of the tumour were also recorded (Table-1). The study protocol received the prior approval of the local ethics committee.

**Table 1.**
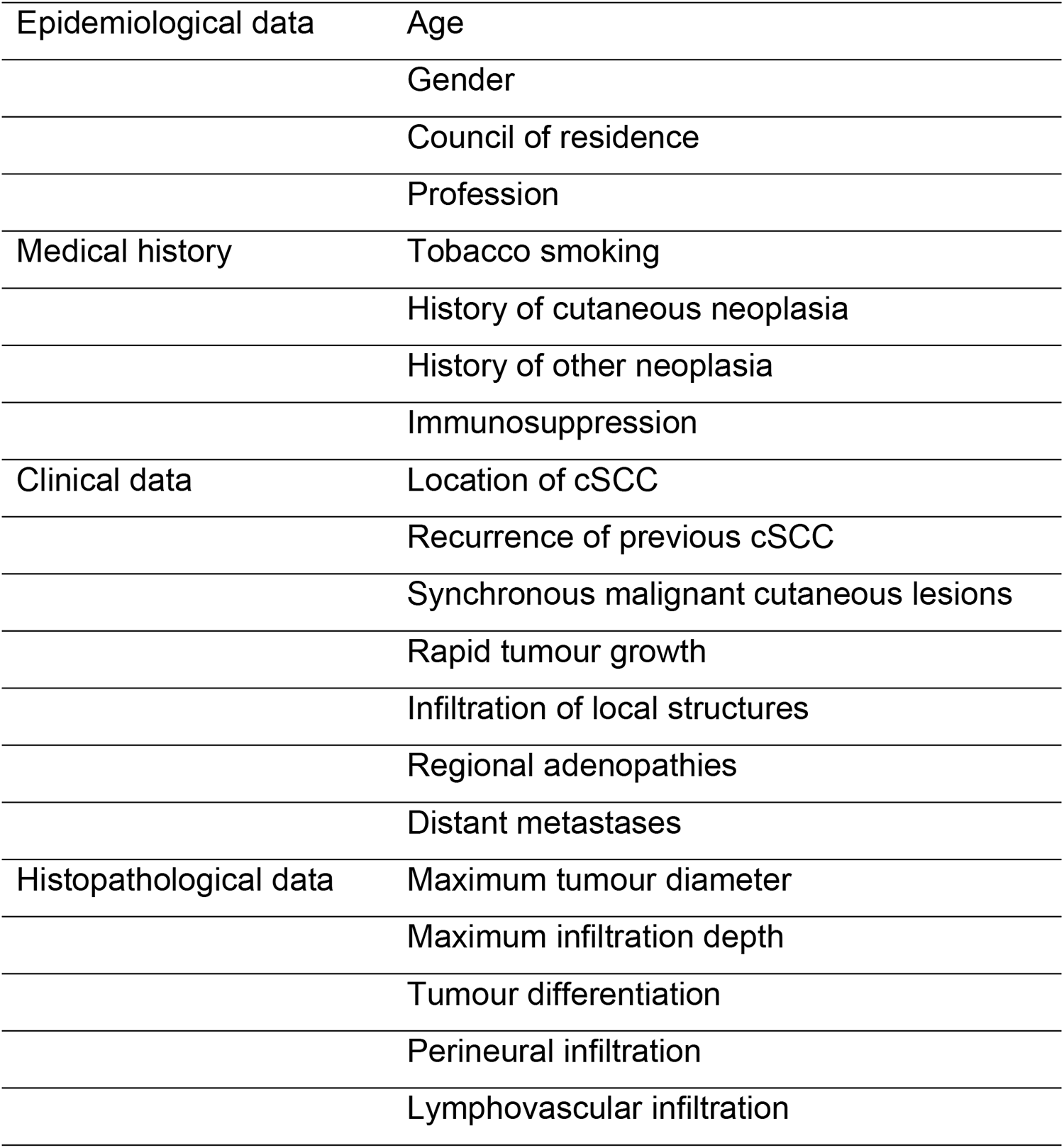
Data collection protocol.

### Calculation of incidence, hours of sunlight per day, environmental levels of radon and natural terrestrial gamma radiation emissions

In order to calculate the incidence of cSCC during the study period, the demographic data for the local councils studied published by the Spanish National Statistics Institute (Instituto Nacional de Estadística) for the year 2020 were taken into account^13^.

Hours of sunlight in the councils studied were obtained from the tables for the year 2020 provided by Climate Data^14^.

Geometric means of environmental radon concentrations for the studied councils were obtained from the open-access data provided by the Galician Radon Laboratory (Laboratorio de Radón de Galicia) of the University of Santiago de Compostela (Spain)^15^.

Natural gamma radiation means were obtained from the MARNA project, carried out by the Spanish Nuclear Safety Council (Consejo de Seguridad Nuclear)^16^.

### Statistical study

The data were collated on a spreadsheet (Microsoft Excel Office 365) and processed with IBM SPSS statistics 22.0 (IBM Corp., Armonk, New York, U.S.A.). The Kolmogorov-Smirnov goodness-of-fit test was employed to assess the normality of the distributions. The study of the correlations between the different variables was carried out using the Pearson test for parametric distributions and the Spearman test in the case of non-parametric distributions or if qualitative variables were used. Multiple linear regression analysis was employed to study the influence of the different variables on the incidence of cSCC.

## Results

A total of 284 patients (165 males and 119 females) diagnosed with cSCC during the 26-month observation period were included in the study. The mean age at tumour diagnosis was 83.15 (8.44) years (81.55 (8.43) years in males and 85.38 (7.96) years in females). The overall incidence was 37.33 cases/100,000 people-year, with a maximum of 131.96 cases/100,000 people-year and a minimum of 4.95 cases/100,000 people-year (Figure 1). The incidence was 39.38 cases/100,000 people-year in males and 34.46 cases/100,000 people-year in females.

**Figure 1.**
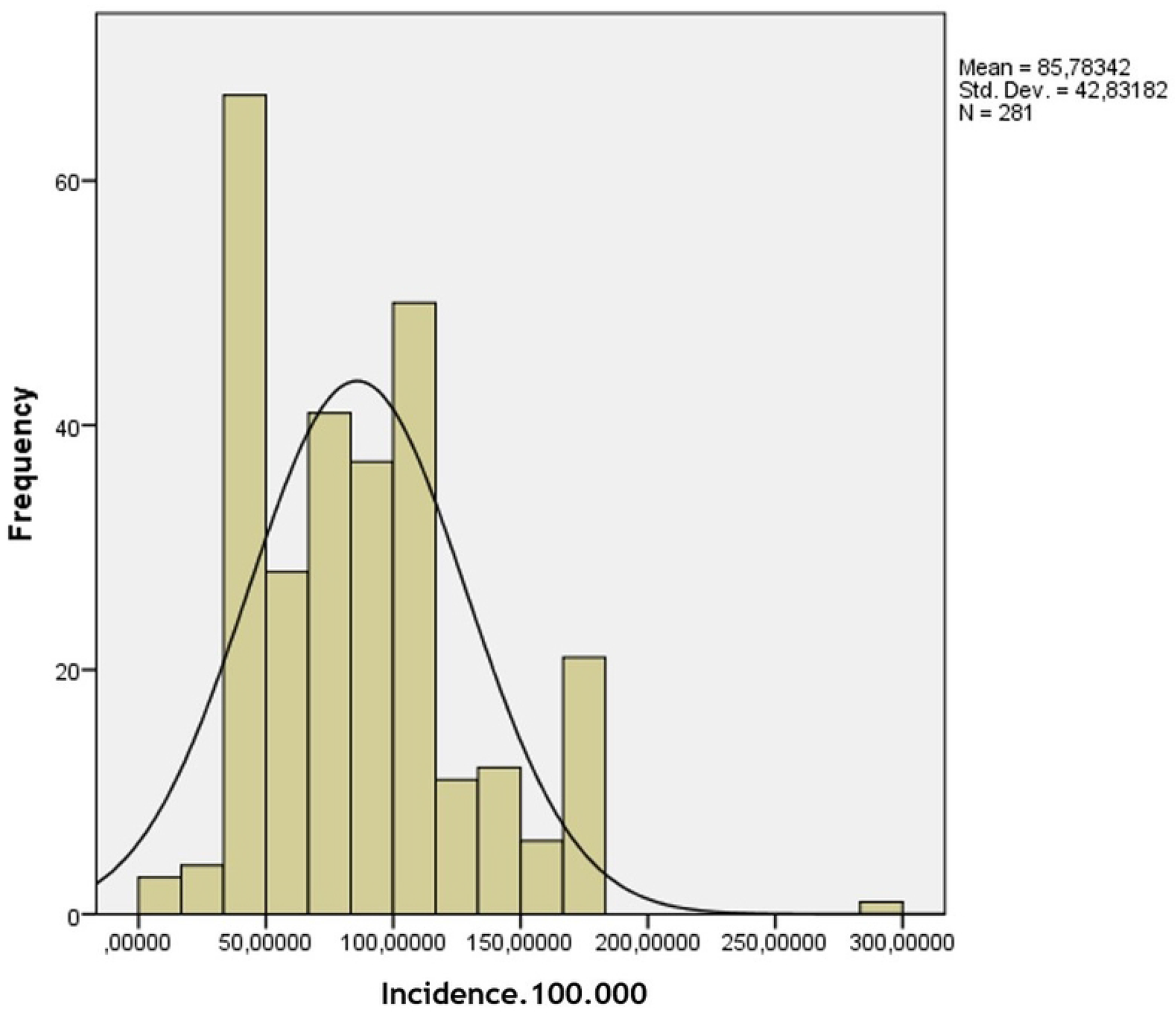

**Figure 2.**
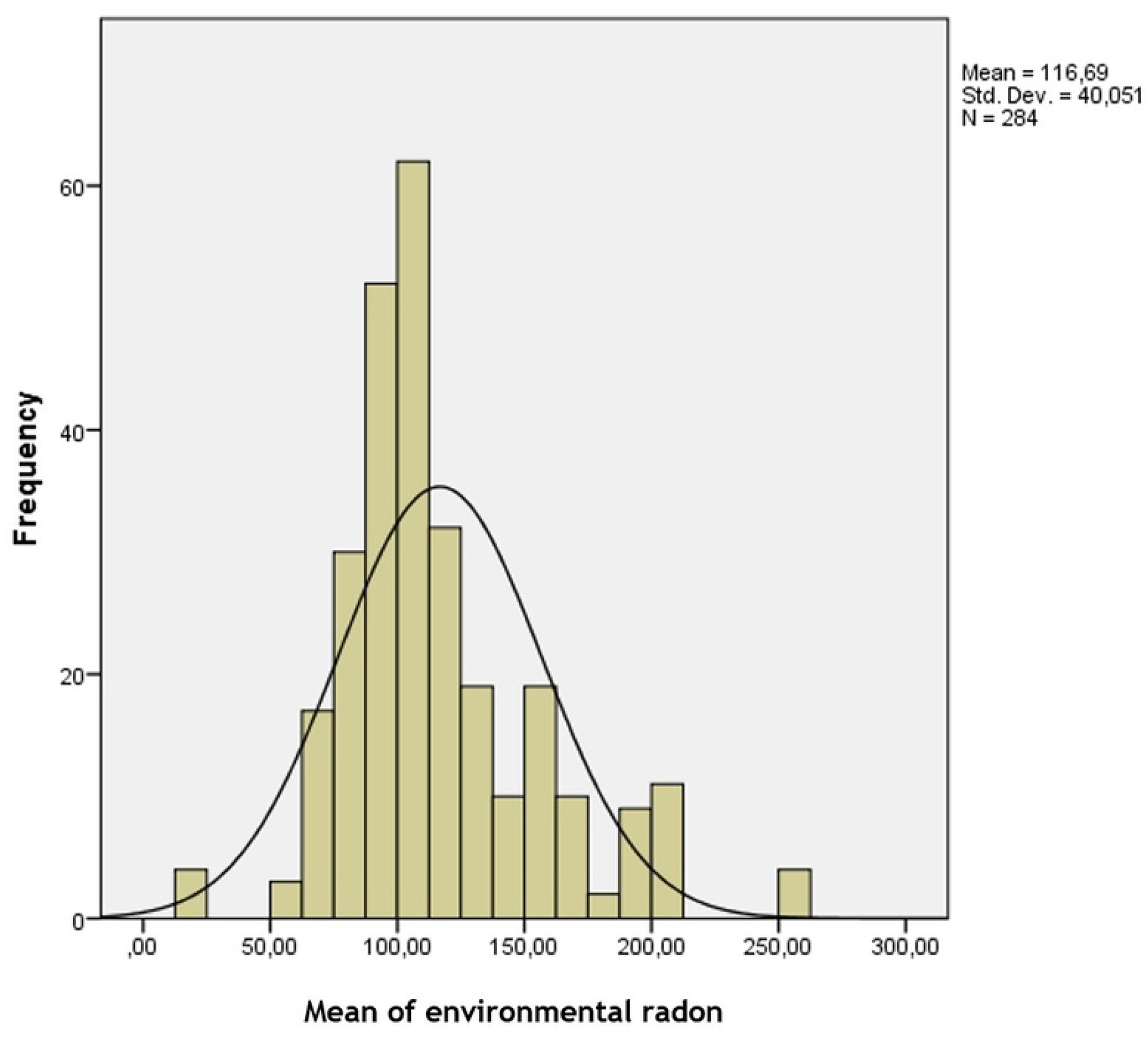

The average hours of sunlight per year in the studied area was 2350.23 hours (185.06), with a maximum of 2717.45 hours and a minimum of 2020.00 hours.

A total of 62.3% (177) of the patients had been employed in professions with a preference for outdoor activity (Table-2). It was not possible to establish the profession of the patient in 4.6% (13) of the cases or whether the activity was mainly carried out indoors or outdoors.

**Table 2.**
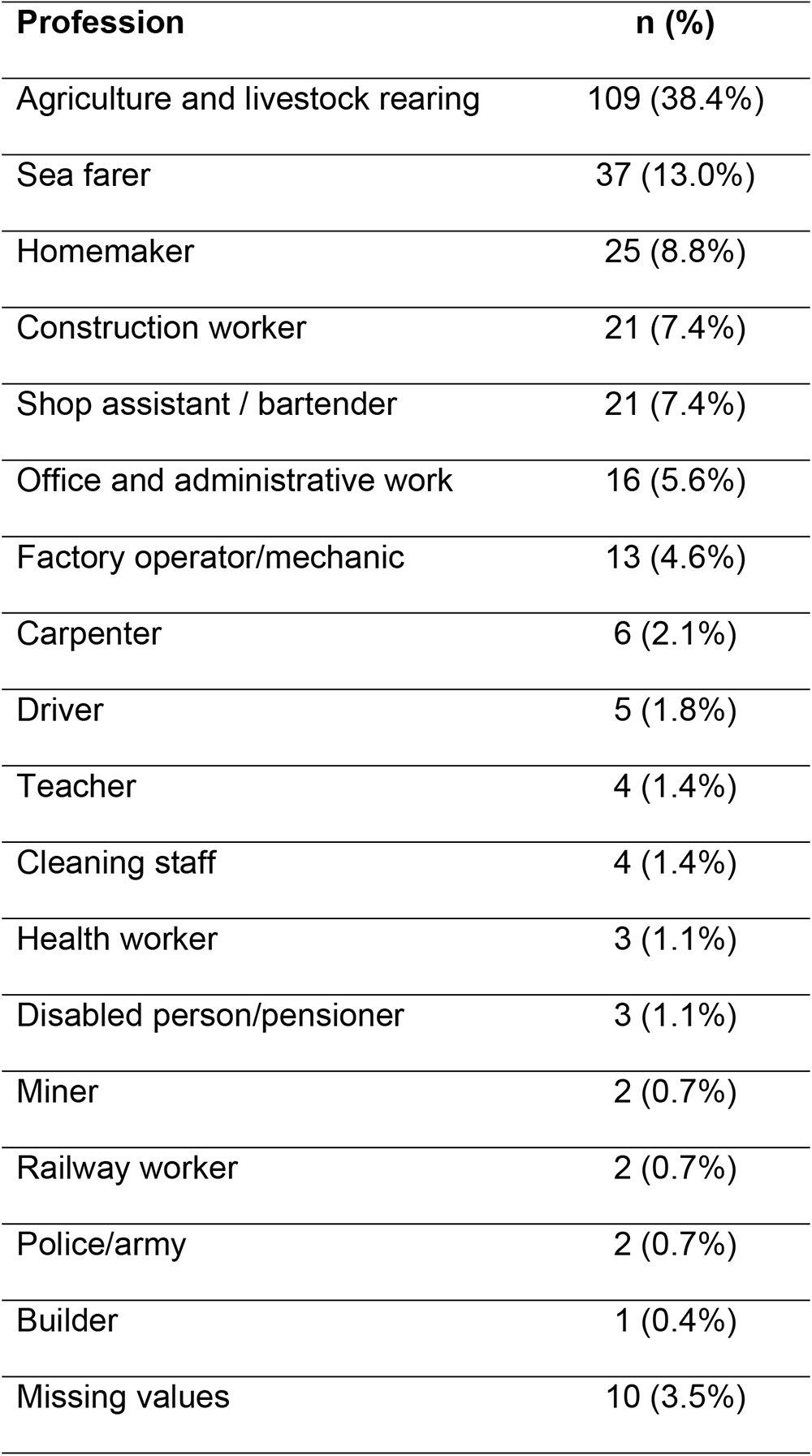
Professions of patients included in the study.

Only 4.9% (14) of the patients were active smokers at the time of cSCC diagnosis, 23.6% (67) were former smokers and 59.5% (169) had never smoked. It was not possible to firmly establish this factor for 12% (34) of the patients included in the study.

No previous diagnosis of a non-cutaneous malignancy had been made in 27.1%(77) of the patients included in the study. The most frequent tumours were prostate adenocarcinoma (17 cases) and rectosigmoid cancer (10 cases). In 30.3% (86) of the cases, a previous diagnosis of other malignant skin lesions had been established. In 25.0% (71) of the patients, there was another synchronous malignant skin lesion at the time of diagnosis. In 52.11% (37) of these cases, the tumours were BCCs, 42.25% (30) of the patients had other cSCCs which were smaller or located in an area other than the head and neck, 4.22% (3) had basosquamous carcinomas and, in one case (1.40%), the synchronous tumour was a lentigo maligna melanoma. In 83.09% (59) of the cases, the synchronous tumour was diagnosed in the skin of the head and neck area.

Immunosuppression was present in 10.9% (31) of the patients at the time of cSCC diagnosis, either secondary to chronic steroid treatment or to an advanced oncological process.

The most frequent location of the tumours was the cheek area (54; 19.0%), followed by the frontotemporal region (51; 17.9%) and the scalp (43; 15.1%) (Table-3).

**Table 3.** Location of cSCC.

| Primary tumour location | n (%) |
| --- | --- |
| Cheek | 54 (19.0%) |
| Frontotemporal region | 51 (17.9%) |
| Scalp | 43 (15.1%) |
| Lower lip | 34 (12.0%) |
| Ear | 28 (9.9%) |
| Nose | 21 (7.4%) |
| Preauricular | 19 (6.7%) |
| Mandibular region | 11 (3.9%) |
| Retroauricular | 9 (3.2%) |
| Upper lip | 5 (1.8%) |
| Neck | 5 (1.8%) |
| Eyelid and orbital region | 4 (1.4%) |

The mean of environmental radon concentration for the patients included in the study according to their council of residence was 116.69 Bq/m^3^ (40.05), with a minimum of 23.58 Bq/m^3^ and a maximum of 257.64 Bq/m^3^ (Figure-2). The mean of natural terrestrial gamma radiation emitted according to their council of residence was 14.25 μRad/hour (3.86), with a maximum of 22 μRad/hour and a minimum of 7 μRad/hour (Figure 3).

**Figure 3.**
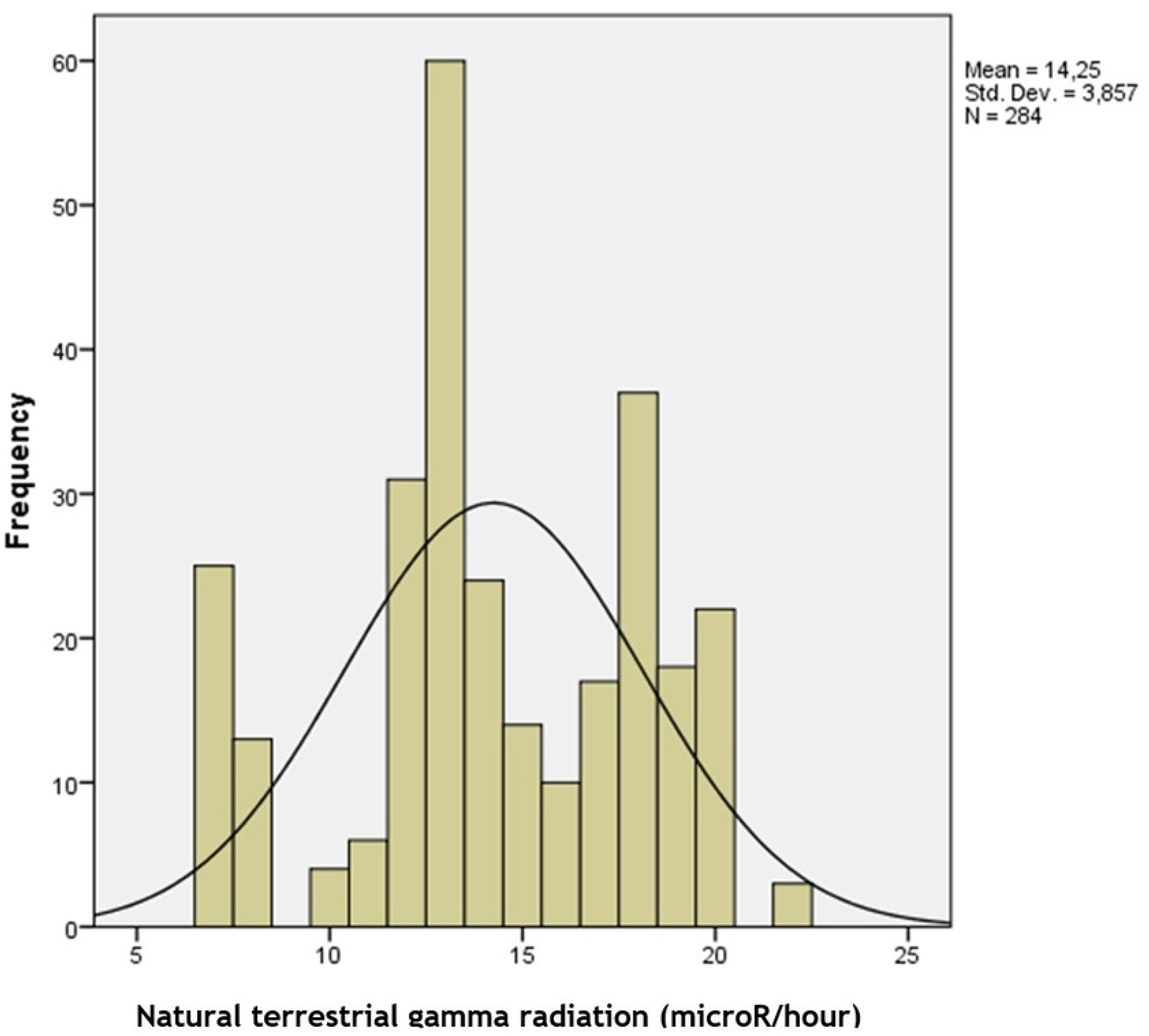

The correlation analysis between the incidence of cSCC and the different variables revealed that there was a statistically significant relationship between cSCC incidence and mean levels of environmental radon in the council of residence (P = 0.02), the mean natural terrestrial gamma radiation emissions in the council of residence (P < 0.05), carrying out an outdoor profession (P < 0.05) and the mean number of hours of sunlight per year in the council of residence (P < 0.05). The multiple linear regression analysis finally excluded environmental radon levels from the model (P = 0.62). Therefore, in the final model, only mean natural terrestrial gamma radiation emissions in the council of residence (P < 0.05), carrying out an outdoor profession (P < 0.05) and the mean number of hours of sunlight per year in the council of residence (P = 0.03) were found to have statistical significance on the incidence of head and neck cSCC (Figure 4).

**Figure 4.**
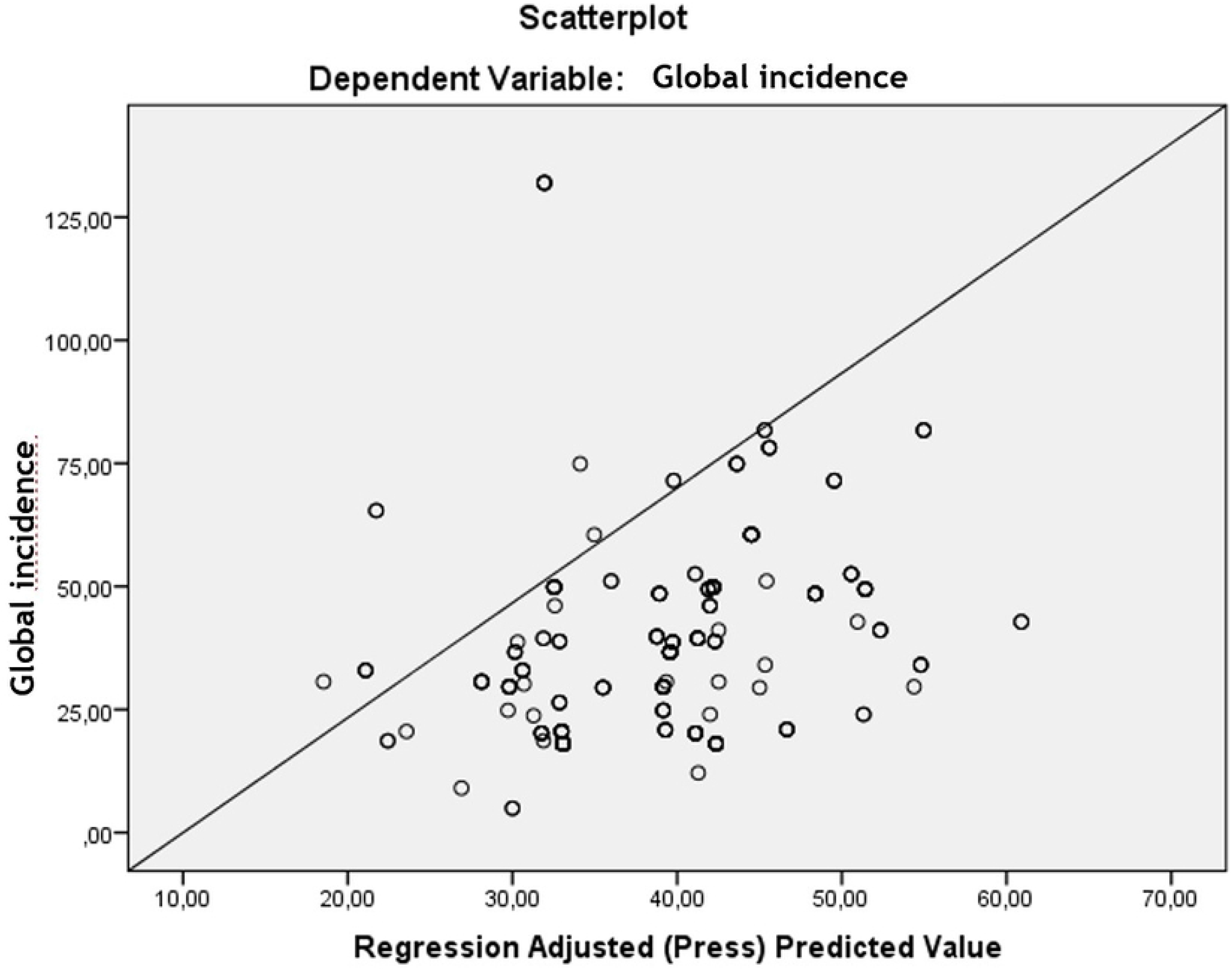

As far as the remaining clinical and histopathological variables included in the study protocol are concerned, in no case was it possible to establish a statistically significant relationship with the mean level of environmental radon, mean natural terrestrial gamma radiation emissions, the mean number of hours of sunlight per year in the council of residence or with occupational solar exposure (Table-4).

Taking the indoor EURATOM safety level for natural gamma radiation emissions of 1 mSv/year (11.41 μRad/hour)^17^ as a reference, 83.10% (236) of the patients included in our study resided in councils in which natural terrestrial gamma radiation emissions were higher than the recommended levels. The incidence of head and neck cSCC in these councils was 39.60 cases/100,000 people-year, whereas the incidence in councils with natural terrestrial gamma radiation emissions below the recommended EURATOM levels was 39.46 cases/100,000 people-year (Figure 5). Although no statistically significant difference can be observed between the two groups, the P value is extremely close to statistical significance (P = 0.05). The Relative Risk (RR) of populations living in councils above the recommended EURATOM values was 1.00.

**Figure 5.**
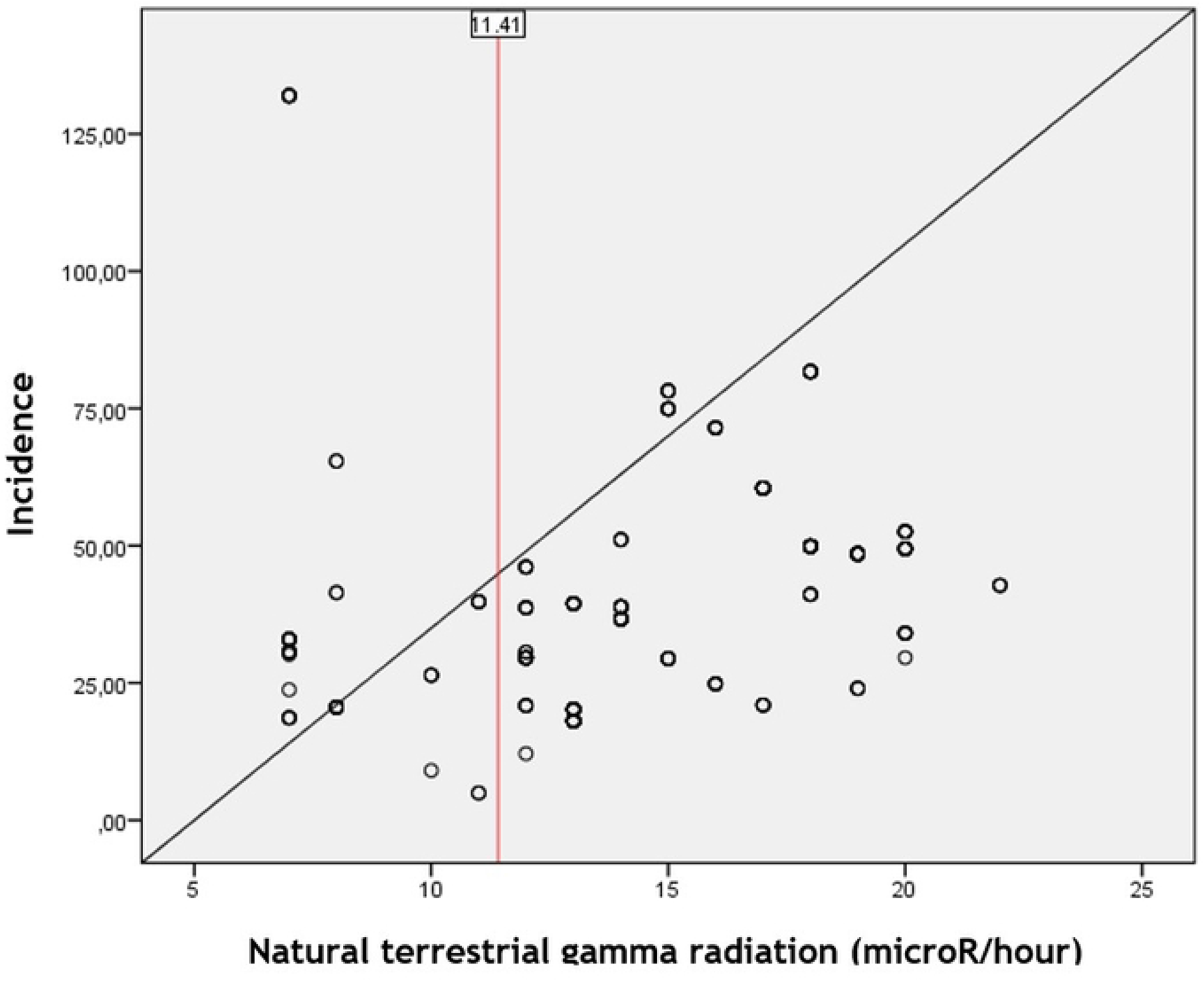
Graphic representation of natural gamma radiation emissions and head and neck SSCC incidence. The EURATOM recommended level of gamma radiation is indicated, showing that slightly more than 80% of the measurements are above the recommended values.

## Discussion

The incidence of head and neck cSCC in our study (37.33 cases/100,000 people-year) shows very similar values to those obtained from the meta-analysis published by Tejera-Vaquerizo (38.16 cases/100,000 people-year), the most exhaustive study carried out to date in Spain in an attempt to determine the incidence of different skin tumours^18^. It is striking that the incidence is similar when solar exposure, the main risk factor in the development of cSCC, is remarkably lower in our series (2350.23 hours of sun/year) than the average values for the country as a whole (2500 hours/year) or in comparison with southern (Seville: 3526 hours/year, Córdoba: 3316 hours/year) or Mediterranean regions (Alicante: 3397 hours/year, Murcia: 3348 hours/year, Mallorca: 3098 hours/year). Based on the results obtained from our study, carried out in a limited area in the northwest of the Iberian Peninsula with an oceanic climate, it can be concluded that the low values of solar exposure observed in comparison with the rest of the country must be compensated by another factor to obtain similar values of head and neck cSCC incidence.

The role that environmental radon plays in the development of certain tumours, as in the case of lung cancer, has already been clearly demonstrated. Indeed, it is now considered to be the first cause of lung cancer among non-smokers^19^. High levels of environmental radon have also been linked to an increased incidence of stomach cancer and related mortality^20–22^, brain and central nervous system tumours^8, 23^ and leukaemia^24^.

The most stable and frequent isotope in the natural environment is ^222^Rn, which results from the decay of ^238^U and has a half-life of 3.824 days. Radon and its progeny, especially short half-life radioisotopes such as ^218^Po and ^214^Po, continue their decay through the emission of alpha particles^25^. Alpha particles have a short range of absorption and low capacity to penetrate the outer layers of the skin. Thus, alpha particles cannot easily reach the basal cell layer of the epidermis, where a theoretical carcinogenic effect would take place. This is the main limiting factor in models which seek to relate environmental radon to the development of cSCC. For this reason, the possible aetiological relationship between radon and tumour locations other than the lung, as well as the pathogenic mechanism, are controversial^26^. To our knowledge, only the study carried out by Wheeler et al. in southwest England has found a significant association between an increased incidence of cSCC and high levels of environmental radon. These results were sustained after adjustment for potential confounders^27^. Other studies have found an increase in the overall incidence of non-melanoma skin cancers^6^, although not specifically of cSCC, in areas with environmental radon levels above 100 Bq/m^3^ when compared to areas with average radon levels below 60 Bq/m^3^. Other studies have found an increase in the incidence of BCC but not of cSCC^8^. An interesting study analysing mortality and cancer incidence among uranium miners exposed to low and moderate doses of environmental radon in the Czech Republic between 1977 and 1992 showed that real incidence of non-melanoma skin cancers was lower than the expected incidence^29^.

The geographical area in which our study was carried out is characterised by the existence of levels of environmental radon above the recommended values in many locations^29^. An association has been demonstrated with an increase in lung^30^, oesophageal^31^ and brain cancer mortality^32^. The mean value for the local councils included in our study was 116.69 Bq/m^3^, which falls within the risk group as defined by Etherington^6^. Safety or action values established by different governments and agencies are usually established between 200 and 400 Bq/m^3^, although it is known that the risk of developing lung cancer has a linear growth with long-term exposure to environmental radon, with no evidence of a defined threshold level. This increase in incidence has been found to be statistically significant even for average values below 200 Bq/m^3 33^. In our study, the correlation of raw data established some degree of statistical significance between environmental levels of radon and incidence of head and neck cSCC. However, in the end, the multiple linear regression model excluded environmental levels of radon as a factor influencing the incidence of cSCC.

Although it has never been postulated as a specific cause of the development of cutaneous neoplasms, we believe that exposure to high levels of natural terrestrial gamma radiation emissions could have an influence on our study population^34^. Natural radionuclides, especially ^40^K from soil and food, are the most frequent source of gamma radiation exposure^35^. Other radionuclides frequently found in nature, depending on the geographical area, are ^238^U, ^235^U and ^232^Th and their progeny.

In the early days of therapeutic ionising radiation, the development of cSCC on the unprotected extremities of technicians and workers exposed to radiation was described^36^, along with an excess in overall mortality due to skin cancer^37^.

Exposure to high-dose gamma radiation may have an influence on the development of cSCC^38^. In the case of exposure to moderate doses, there appears to be a clear impact on the development of BCC, although this is not so clear in the case of cSCC^39–41^. Lichter et al. demonstrated an increased risk of cSCC in patients who had undergone previous ionizing radiation therapy due to conditions other than skin cancer. This increased incidence proved close to statistical significance. The *odds ratio* for cSCC seemed to increase with the frequency and number of treatments carried out^3^. In the case of low-dose gamma radiation, Daniels RD et al. linked prolonged exposure to a significant increase in leukaemia diagnoses^42^. However, it was not until the study carried out by Azizova et al. in 2021 that a significant relationship was demonstrated between exposure to low-dose ionising radiation for extended periods of time and an increase in the incidence of skin tumours^43^. This study, carried out in the Chelyabinsk region of the Russian Federation among nuclear industry workers, found that the incidence of non-melanoma skin neoplasms was significantly associated with cumulative external gamma dose. Relative risk was found to be significantly increased for both cSCC and BCC. This exposure to low doses of gamma radiation over long periods of time is the most similar scenario to that presented in our study, taking into account the fact that no previous study has ever considered only natural background radiation in order to discover its influence on cutaneous malignancies.

## Conclusion

In our study, emissions of natural terrestrial gamma radiation, together with hours of sunlight and predominantly outdoor professions, are the main causes of the development of cSCC. To date and to the best of our knowledge, emissions of natural terrestrial gamma radiation have never been proposed as a factor having an influence on the development of cutaneous neoplasms. The implications that this could have in areas with naturally high terrestrial gamma radiation emissions, especially in geographical areas in which the classical studied risk factors for development of cSCC are not so present, should be more exhaustively studied in order to assess the true weight of this factor.

## Data Availability

Data cannot be shared publicly because of we are working with private clinical features obtained from medical records.

## Acknowledgments

We would like to thank Mr Paul Jonathan Lacey B.A. for his kind willingness to proofread the English manuscript.

